# Approximating R1 and R2: a quantitative approach to clinical weighted MRI

**DOI:** 10.1101/2024.05.22.24307730

**Authors:** Shachar Moskovich, Oshrat Shtangel, Aviv A. Mezer

**Affiliations:** The Hebrew University of Jerusalem, Israel

## Abstract

Weighted MRI images are widely used in clinical as well as open-source neuroimaging databases. Weighted images such as T1-weighted, T2-weighted, and proton density-weighted (T1w, T2w, and PDw, respectively) are used for evaluating the brain’s macrostructure; however, their values cannot be used for microstructural analysis, since they lack physical meaning. Quantitative MRI (qMRI) relaxation rate parameters (e.g., R1 and R2), and related relaxivity coefficients, do contain microstructural physical meaning.

Nevertheless, qMRI is rarely done in large-scale clinical databases.

Currently, the weighted images ratio T1w/T2w is used as a quantifier to approximate the brain’s microstructure. In this paper, we propose three additional quantifiers that approximate quantitative maps, which can help bring quantitative MRI to the clinic for easy use.

Following the signal equations and using simple mathematical operations, we combine the T1w, T2w, and PDw images to estimate the R1 and R2.

We find that two of these quantifiers (T1w/PDw and T1w/ln(T2w)) can serve as a semi-quantitative proxy for R1, and that (ln(T2w/PDw)) can approximate R2.

We find that this approach also can be applied to T2w scans taken from widely available DTI datasets. We tested these quantifiers on both *in vitro* phantom and *in vivo* human datasets. We found that the quantifiers accurately represent the quantitative parameters across datasets. Finally, we tested the quantifiers within a clinical context, and found that they retain tissue information across datasets. Our work provides a simple pipeline to enhance the usability and quantitative accuracy of MRI weighted images.

## Introduction

Quantitative MRI (qMRI) is a powerful tool for studying the brain’s microstructure. It can be used to produce parametric maps, which allow for reliable comparisons across subjects, time points, and MRI scanners^1^. Recent studies have proposed that qMRI, when combined with appropriate biophysical models, can be used to obtain *in vivo* histological information about the human brain^2,3^. Quantitative relaxometry maps such as R1 (1/T1) and R2 (1/T2) are sensitive to myelin content, iron concentration, and water content^4–7^. Another quantitative map is Proton Density (PD), which is sensitive to the brain’s water content^8^.

While there are clear advantages in using biophysically meaningful quantitative MRI, it is considered to be technically challenging and requires long scanning times, which means it is rarely used in clinical settings. On the other hand, clinical MRI assessment requires relatively short scanning times, and typically rely on weighted images, which are not suited for quantitative biophysical analysis of tissue microstructure. Additionally, many large open-source datasets of neurological disorders (e.g., PPMI (https://www.ppmi-info.org/access-data-specimens/download-data), ADNI (https://adni.loni.usc.edu/), UK-Biobank (https://www.ukbiobank.ac.uk/) and ABCD (https://abcdstudy.org/)) contain weighted images as well as diffusion tensor imaging (DTI) data. Therefore, there would be a great benefit in linking the widely available clinical MRI weighted data and open-source databases to known qMRI parameters (e.g., R1 and R2).

In recent years, many studies have utilized the ratio of T1-weighted and T2-weighted images (T1w/T2w) as a semi-quantitative measure. On one hand, it has some quantitative properties, as it minimizes the instrumental bias. On the other hand, it is non-quantitative, since it was not fitted using the physical signal equation, nor is it a direct estimate of a physical quantity^10^. Since then, T1w/T2w became widely available, and has been suggested as an indicator of tissue integrity and myelination^11,12^. The distribution of T1w/T2w in the brain has been linked to myelin expression^13–19^ and brain development^12,20–22^. Abnormal changes in T1w/T2w have been associated with neurodegenerative disorders such as Parkinson’s disease^23^, multiple sclerosis^24–26^, Alzheimer’s disease^27^, and Huntington’s^28^ disease, and even affective diseases such as schizophrenia^29^.

Another previously suggested method to measure quantitative information using weighted images is the log-linear approach, which utilized gradient-echo sequences containing multiple echo times to quickly measure R2^30^.

In this study, we test two weighted image quantifiers (i.e., semi-quantitative combinations of weighted images) based on three common weighted images (T1w, T2w, and PDw) and using mathematical combinations such as ratios and logarithms. We call these two semi-quantitative qualifiers T1w/PDw and ln(T2w/PDw), and we show that they can approximate the observed, single-exponent qMRI parameters R1 and R2, respectively. It is a common practice in the qMRI field to represent the tissue relaxations using a single exponential rate constant^1,2,30–33^ . However, biological tissue is ideally described using multi-exponential relaxation signal equations, since they can better account for the multi-compartmental organization of the tissue. Naturally, using weighted images, we aim to approximate the single-exponent representation. We validate our results both on a controlled lipid phantom system and on two human datasets. Furthermore, we modify the existing T1w/T2w quantifier and propose a third quantifier (T1w/ln(T2w)), which can better approximate R1.

Finally, we apply our analyses to a clinical dataset from the PPMI, compare it with our previous results, and find the quantifiers to be robust across datasets. In summary, we propose a way to utilize weighted images, whose common use is clinical, to approximate quantitative MRI parametrization.

## Results

In the following sections, we first test the theory introduced in Methods section on synthetic phantom data, and produce two semi-quantitative maps. Next, we test the theory *in vitro* using lipid phantoms, and *in vivo* using two human datasets.

### Quantitative MRI values can be approximated using weighted images

We created simulated data for R1 and R2 values (which varied within the common range of R1 and R2 values), and used them in Eq. 1-4 to simulate semi-quantitative T1w/PDw and ln(T2w/PDw).

As expected from Eq. 3 and 4, we produced perfect linear relationships between T1w/PDw and the R1 map, as well as a perfect relationship between ln(T2w/PDw) and R2 map (*R*^2^ = 1; Fig. 1). These results indicate that weighted images theoretically can provide a perfect estimate of the single-exponent, observed quantitative maps in a noise-free environment.

**Fig. 1.**
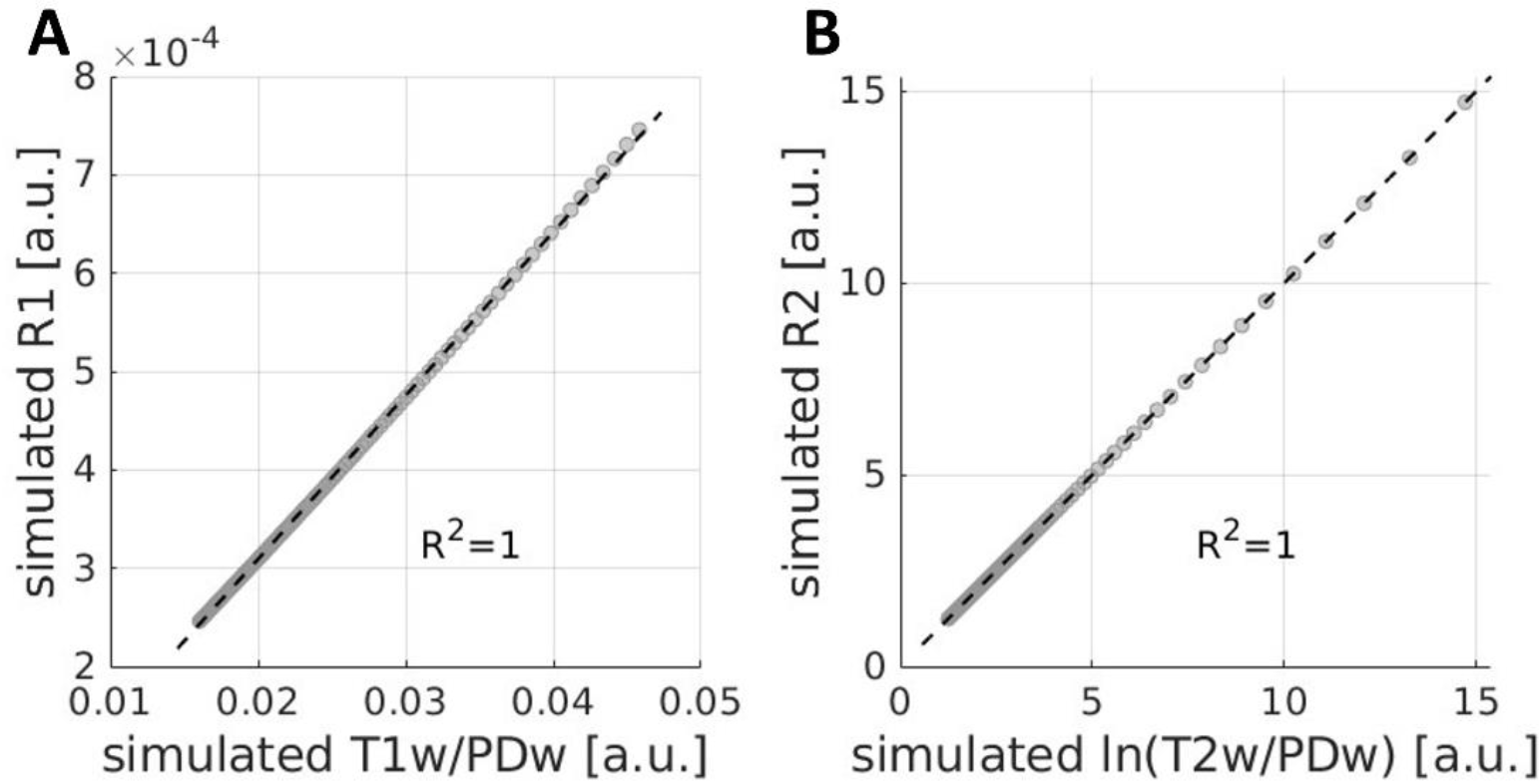
The simulated T1w/PDw and ln(T2w/PDw) quantifiers yield a perfect correlation with simulated quantitative maps R1 and R2. As predicted from the MR relaxation equations, the weighted images quantifiers are in perfect agreement with the quantitative relaxation rate. Simulated values were calculated from the single-exponent relaxation rates produced between the common ranges of R1 and R2. Correlations are shown for **(A)** simulated R1 and simulated T1w/PDw; and (**B)** simulated R2 and simulated ln(T2w/PDw). Data points represent simulated values. The black dashed line is the regression line. a.u. – arbitrary units.

To validate the utility of the semi-quantitative quantifiers *in vitro*, we first tested it using phantom data. Specifically, we evaluated the correlation of R1 with T1w, T1w/T2w, and T1w/PDw. We found significant correlations for all three comparisons (Fig. 2A-C). T1w/T2w showed the weakest correlation, and T1w/PDw showed the strongest. We performed 1,000-fold cross-validation (CV) for each pair of variables: each time, we trained a linear model on 90% of the voxels and tested it on the remaining 10%, and then calculated the RMSE. We found that T1w/PDw had the lowest mean RMSE (0.0734 s^-1^), compared to T1w (0.102 s^-1^) and T1w/T2w (0.105 s^-1^). Similarly, we calculated the correlations of R2 with T2w, T1w/T2w, and ln(T2w/PDw). We found that the strongest correlation was with ln(T2w/PDw) (Fig. 2D-F). We did the same CV analysis, and found the lowest error with ln(T2w/PDw) (0.0003 s^-1^), compared to R2w (0.002 s^-1^) and T1w/T2w (0.002 s^-1^). These findings highlight the benefit of the T1w/PDw and ln(T2w/PDw) quantifiers, as they robustly approximate quantitative relaxation values in phantoms.

**Fig. 2.**
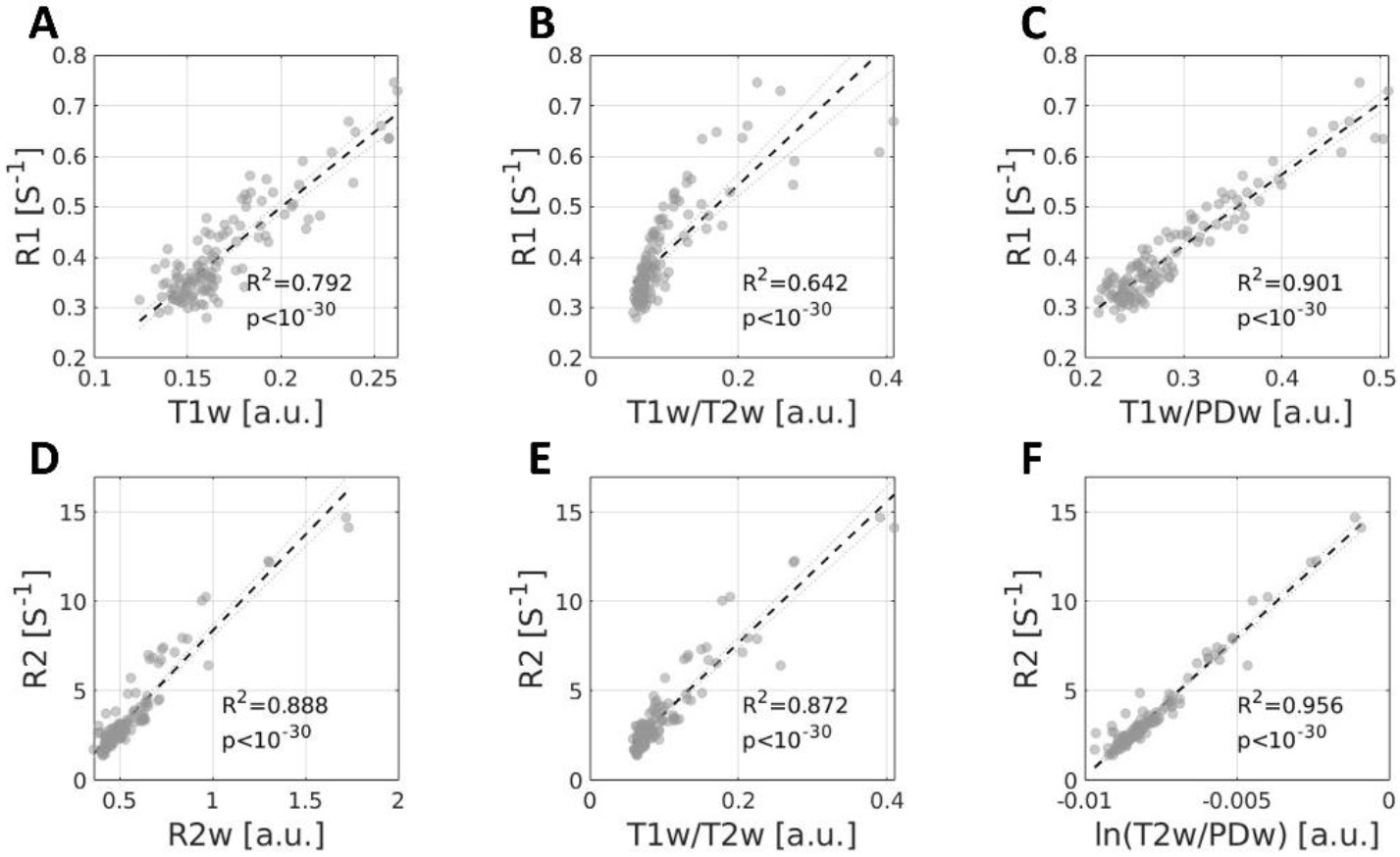
R1 and R2 can be approximated using the T1w/PDw and ln(T2w/PDw) quantifiers in lipid phantom data. A comparison of R1 and R2 to different quantifiers derived from weighted images. **(A-C)** Correlation coefficients (R^2^) between the R1 map and: **(A)** T1w; **(B)** T1w/T2w; and **(C)** T1w/PDw. T1w/PDw yields the highest correlation. **(D-F)** Correlation coefficients (R^2^) between the R2 map and: **(D)** R2w; **(E)** T1w/T2w; and **(F)** ln(T2w/PDw). ln(T2w/PDw) yields the highest correlation. Data is taken from phantom scans (Dataset A), and datapoints represent the phantom samples’ values. The black dashed lines represent the regression line. a.u. – arbitrary units.

To test the applicability of the *in vitro* results on *in vivo* human brain data, we used a human dataset of 22 subjects (Dataset B). We pooled whole-brain images from all subjects, and calculated the correlations of the weighted images T1w, T1w/T2w and T1w/PDw with R1. Here we used MPRAGE images instead of SPGR-based images for T1w, as MPRAGE is the T1-weighted image most commonly employed. (Additionally, we assume that both MPRAGE and SPGR-based T1w images similarly depend on T1 and PD contributions; see *Methods*.) As in the phantom data, here also T1w/PDw yielded the highest correlation with R1 (Fig. 3A-C). On the single-subject level, we found that both T1w and T1w/PDw show strong correlations with R1, and that both are significantly higher than for T1w/T2w (see Supplementary Fig. S1A). Together, these results suggest that T1w/PDw is a better approximation for R1 compared to both T1w and T1w/T2w.

**Fig. 3.**
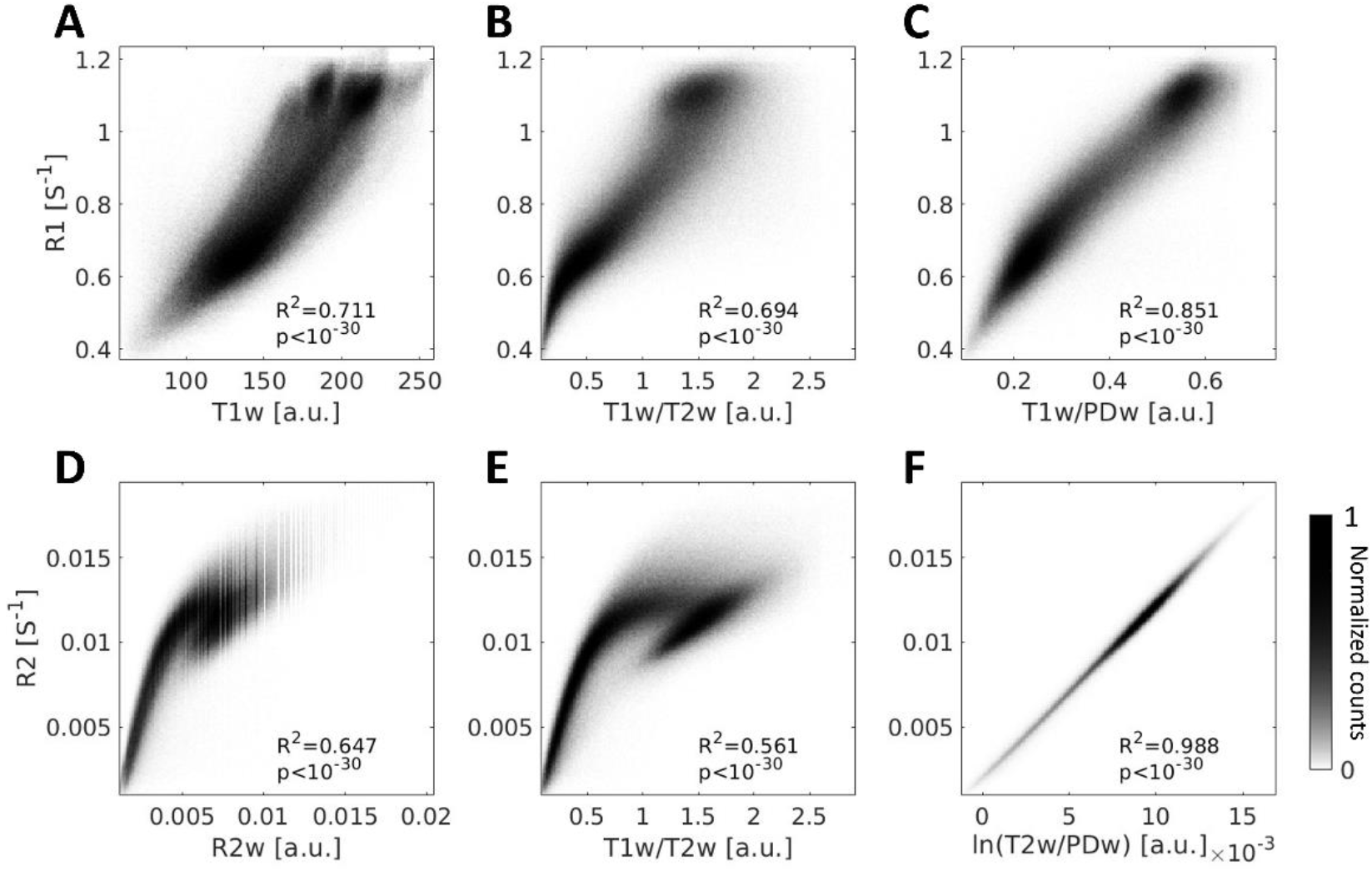
R1 and R2 can be approximated using the T1w/PDw and ln(T2w/PDw) quantifiers in human data *in vivo*. A comparison of R1 and R2 to different quantifiers derived from weighted images. **(A-C)** Voxel-wise 2D-histograms of R1 voxels pooled from Dataset B (N=22) with either **(A)** T1w; **(B)** T1w/T2w; or **(C)** T1w/PDw. The T1w/PDw quantifier shows the highest correlation with R1. **(D-F)** Voxel-wise 2D-histograms of R2 voxels pooled from Dataset B (N=22) with either **(D)** R2w; **(E)** T1w/T2w; or **(F)** ln(T2w/PDw). The ln(T2w/PDw) quantifier shows the highest correlation with R2. R^2^ represents Pearson’s correlation coefficient. Colorbar represents normalized voxel counts.

Similarly, for the R2 approximates, we replicated the phantom results in humans. We found that ln(T2w/PDw) had the highest correlation with R2, among all other tested quantifiers (Fig. 3D-F, also for single-subject in Dataset B see Supplementary Fig. S1D). To confirm that the “*ln”* operation itself was not solely responsible for this effect, we also examined ln(R2w) and ln(T1w/T2w), which showed weaker correlations with R2 than did ln(T2w/PDw) (Fig. 4).

**Fig. 4.**
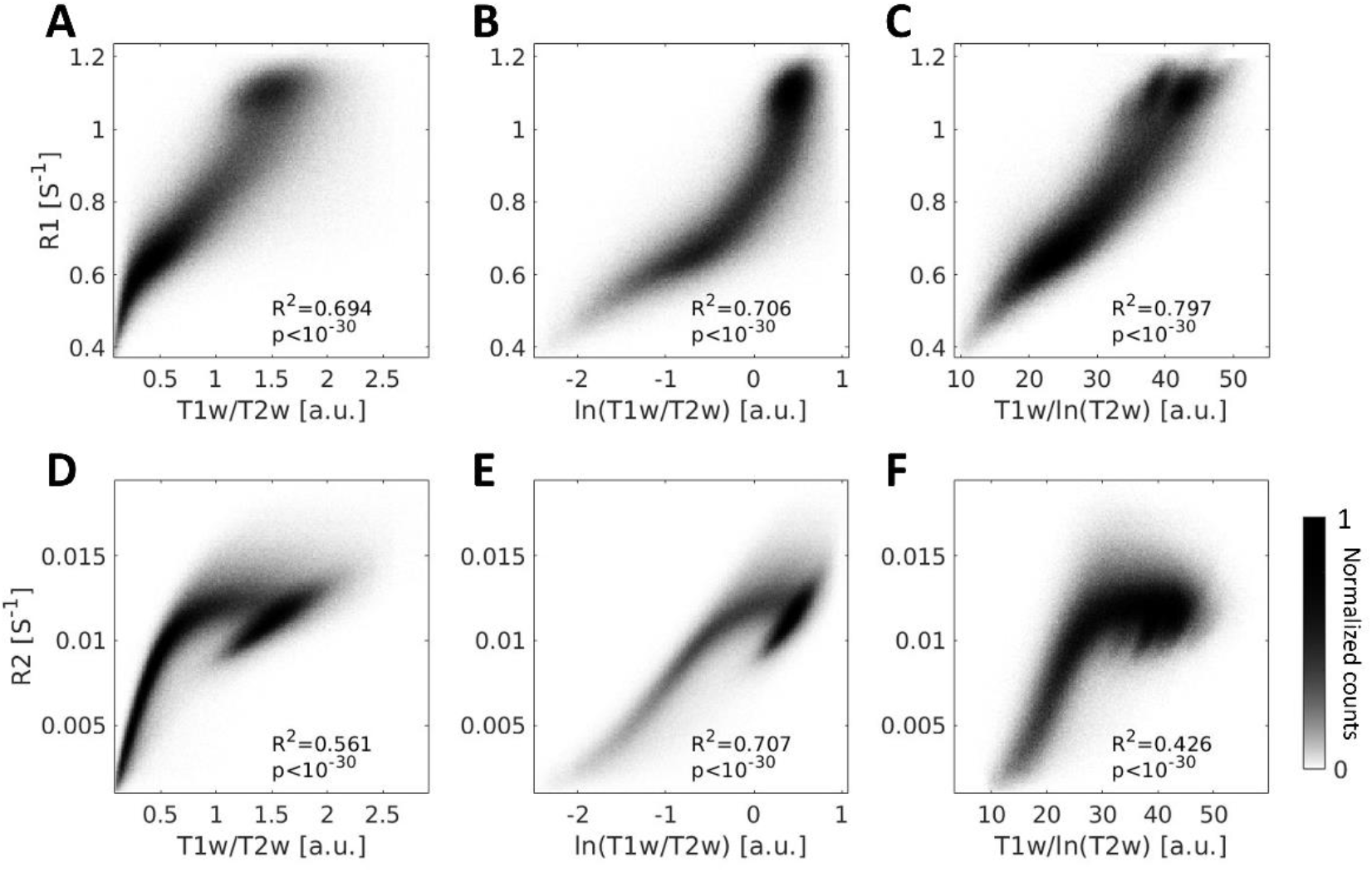
Adjusting the T1w/T2w quantifier can improve the approximation of R1 and R2. A comparison between R1 and R2 and the modified quantifiers that are based on T1w/T2w. **(A-C)** Voxel-wise 2D-histograms of R1 voxels pooled from Dataset B (N=22) with either **(A)** T1w/T2w; **(B)** ln(T1w/T2w); or **(C)** T1w/ln(T2w). T1w/ln(T2w) improves the T1w/T2w linear approximation of R1. **(D-F)** Voxel-wise 2D-histograms of R2 voxels pooled from the same dataset with either **(D)** T1w/T2w; **(E)** ln(T1w/T2w); or **(F)** T1w/ln(T2w). ln(T1w/T2w) improves the T1w/T2w linear approximation of R2. R^2^ represents Pearson’s correlation coefficient. Colorbar represents normalized voxel counts.

### Validating the semi-quantitative contrasts

To test the residual contributions of spatial inhomogeneity biases to the tested quantifiers, we applied a bias correction algorithm^34,35^. This correction accounts for inhomogeneous bias fields that might not cancel out when dividing the weighted images (to produce T1w/PDw and ln(T2w/PDw)). We found similar results to the non-corrected results (Fig. 3) when pooling whole-brain images from all subjects of Dataset B: that is, the highest correlation with R1 was with T1w/PDw, and the highest correlation with R2 was with ln(T2w/PDw). However, in this analysis we detected a separation between subjects (apparent especially in supplementary Fig. S2A). This separation suggests that the bias correction might introduce an additional shift in the range of values of the semi-quantifiers, thus generating another source of variation. To overcome this shift, we calculated the correlations with the quantitative maps R1 and R2 for individual subjects (supplementary Fig. S1B,E). Our results with the individualized correction are similar to those without the correction, suggesting this bias-correction step is not beneficial in our dataset. Additionally, we normalized the values for each subject (see *Methods*), and got similar trends as shown in Fig. 3, where T1w/PDw best approximates R1 and ln(T2w/PDw) best approximates R2 (Supplementary Fig. S3).

In order to rule out that our observations are due to the dependency between commonly acquired images or resampling bias due to difference in images resolution, we scanned an additional independent dataset, such that each weighted image was acquired using a different scanning sequence but at the same resolution (Dataset C). We then pooled whole-brain images from Dataset C’s subjects (N=3), and indeed we replicated the result in Dataset B. In particular, we note our finding that ln(T2w/PDw) is the best approximation of R2 in this dataset as well (Supplementary Fig. S4). However, unlike our results from Dataset B, there was no advantage for T1w/PDw over T1w or T1w/T2w. We suggest this may be related to the small number of subjects in Dataset C.

### Modification of T1w/T2w reveals R1 approximation

T1w/T2w is a commonly used semi-quantitative quantifier^11–17,20–22^. Following our analysis, we ask whether T1w/T2w could more accurately approximate the quantitative maps R1 and R2. Since the PDw image is less common than T1w and T2w throughout clinical datasets, we ask whether we can optimize the utilization of T1 and T2 weighted images. In Figs. 2B,E and 3B,E, we report that the T2w and T1w/T2w images show an exponential change with respect to R2 or R1. Hence, we calculated the quantitative maps’ correlations with ln(T1w/T2w) for humans.

Based on Eq. 2, we suggest applying the natural logarithm on T2w alone, and therefore calculated T1w/ln(T2w) for Dataset B. Interestingly, when pooling whole-brain images from all subjects, we observed high correlations between R1 and ln(T1w/T2w) and between R1 and T1w/ln(T2w) (*R*^2^ = 0.706, *p* < 10^−7^; *R*^2^ = 0.797, *p* < 10^−7^ respectively; Fig. 5A-C). On the single-subject level, R1 showed a high correlation with T1w/ln(T2w), though it was not significantly different from R1’s correlations with T1w and with T1w/PDw (Supplementary Fig. S1A). We replicated this results in Dataset C (Supplementary Fig. S4A-C, and single-subject results in Supplementary Fig. S1C). Hence, we found T1w/ln(T2w) to be a better approximation of R1 than is T1w/T2w. It is worth noticing that any shared bias between the images will not be cancelled when taking T1w/ln(T2w) instead of the other options (i.e., T1w/T2w or ln(T1w/T2w)).

**Fig. 5.**
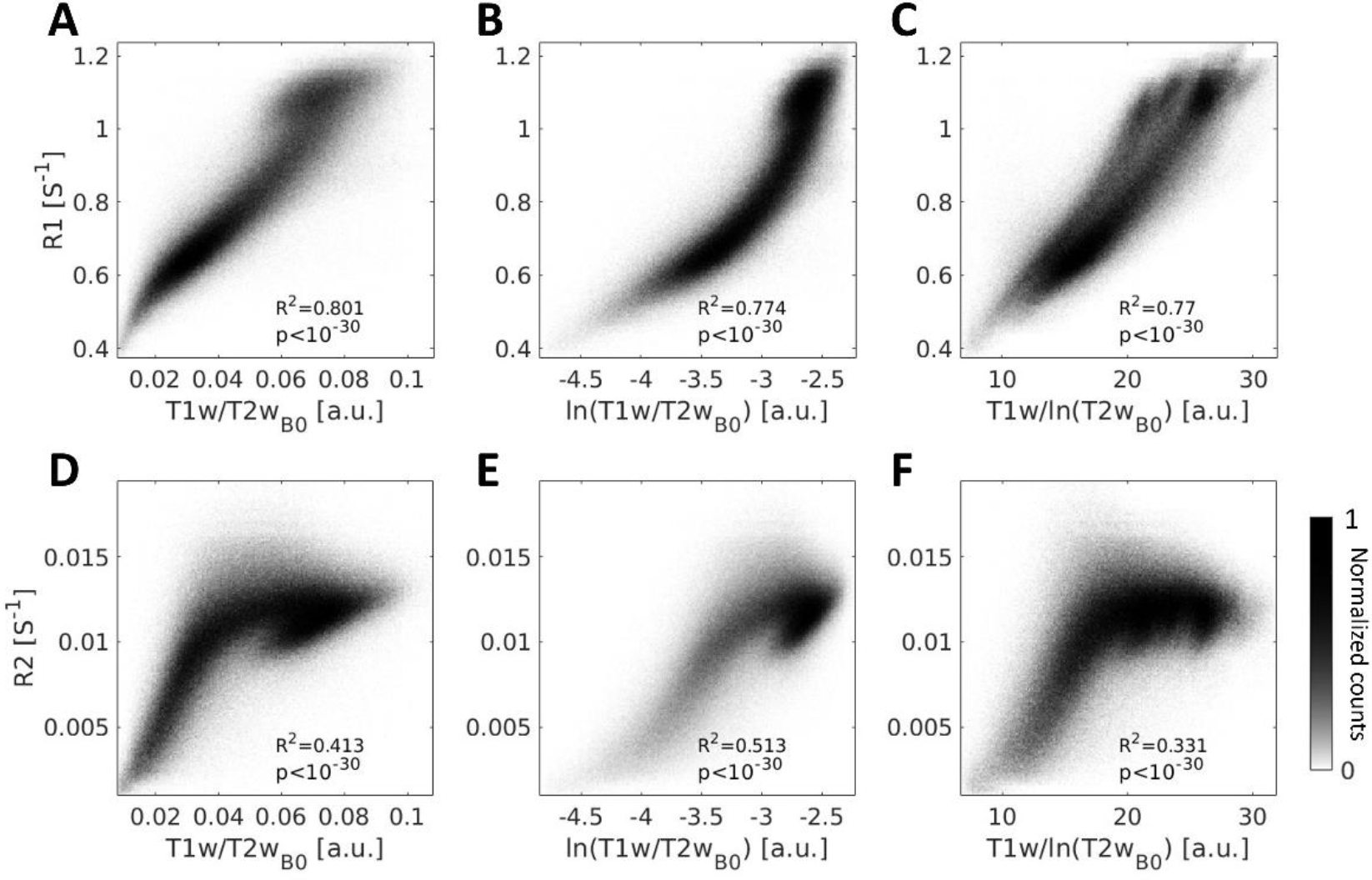
DTI T2w_B0_ can replace T2w for a semi-quantitative quantifiers. A comparison of R1 and R2 to different quantifiers derived from weighted and DTI images, across subjects. **(A-C)** Voxel-wise 2D-histograms of R1 voxels pooled from 20 subjects (Dataset B) with either **(A)** T1w/ T2w_B0_; **(B)** ln(T1w/ T2w_B0_); or **(C)** T1w/ln(T2w_B0_). **(E-F)**, The same as **(A-C)** but for R2 approximation. Group correlations between R2 and different quantifiers across subjects. In this dataset, T2w_B0_ can be used for the quantifiers. R^2^ values represent Pearson’s correlation coefficient. Colorbar represents normalized voxel counts.

Next, we tested the correlations of R2 with each of T1w/T2w, ln(T1w/T2w), and T1w/ln(T2w) in Dataset B. Of these three correlations, we found the highest to be with ln(T1/T2w) (*R*^2^ = 0.706, *p* < 10^−7^, Fig. 5D-F). However, for the single-subject analysis, none of the T1w/T2w variations was as good as ln(T2w/PDw) in approximating R2 (Supplementary Fig. S4D-F, and single-subject results in Supplementary Fig. S1D). This result was replicated in Dataset C (Supplementary Fig. S1F).

### Quantitative R1 can be approximated using DTI data

One main goal of this work is to utilize existing non-quantitative data to extract quantitative information, so we wanted to generalize our analysis to widely available datasets that contain T1w data and DTI data but not T2w data (e.g., UK Biobank). Therefore, we repeated our analysis in Dataset B, using the B0 images (i.e., non-diffusion T2w images, hereafter T2w_B0_) that are collected with every DTI scan. Importantly, T2w_B0_ is a measure taken with an EPI readout, in different from the T1w, and a nonlinear image registration is needed^36,37^. We replicated the main T2w result for T2w_B0_: We compared the correlations between R1 and either T1w/T2w_B0_, T1w/ln(T2w_B0_), and ln(T1w/ T2w_B0_). We found that T1w/ln(T2w_B0_) provides the best approximation of R1(Fig. 5). We also compared the correlations between R2 and these three quantifiers, and found that ln(T1/ T2w_B0_) can approximate R2. Notably, in this dataset, using any quantifier with T2w_B0_ instead of T2w gives a better estimation of R1 and a worse estimation of R2.

### The semi-quantitative quantifiers are robust across datasets

To test the robustness of the proposed structural quantifiers, we analyzed data of healthy subjects from open-source Parkinson’s disease data (PPMI^38^ – Dataset D). We compared between the quantifiers in Dataset D and Dataset B’s quantitative maps R1 and R2. To compare between the datasets, we used the median values of 14 gray- and white-matter ROIs across the brain (Fig. 6). Similar to the result that we found within subjects in Dataset B (Figs. 3 and 4), we also found high correlations between the quantifiers calculated for Dataset D and the quantitative maps calculated for Dataset B. The quantitative R1 map of Dataset B correlated strongly with Dataset D’s T1w/PDw (*R*^2^ = 0.962, *p* < 10^−30^, Fig. 6A) and also with T1w/ln(T2w) (*R*^2^ = 0.961, *p* < 10^−30^, Fig. 6B. We also found a significant correlation between quantitative R2 map of Dataset B and the semi-quantitative ln(T2w/PDw) of Dataset D (*R*^2^ = 0.761, *p* < 0.005, Fig 6C). These high correlations between semi-quantitative quantifiers produced from clinical data and quantitative maps mean that semi-quantitative approximation can be also done using weighted images originated from clinical data. Hence, these results paves the way for future semi-quantitative analysis to be performed in clinical datasets.

**Fig. 6.**
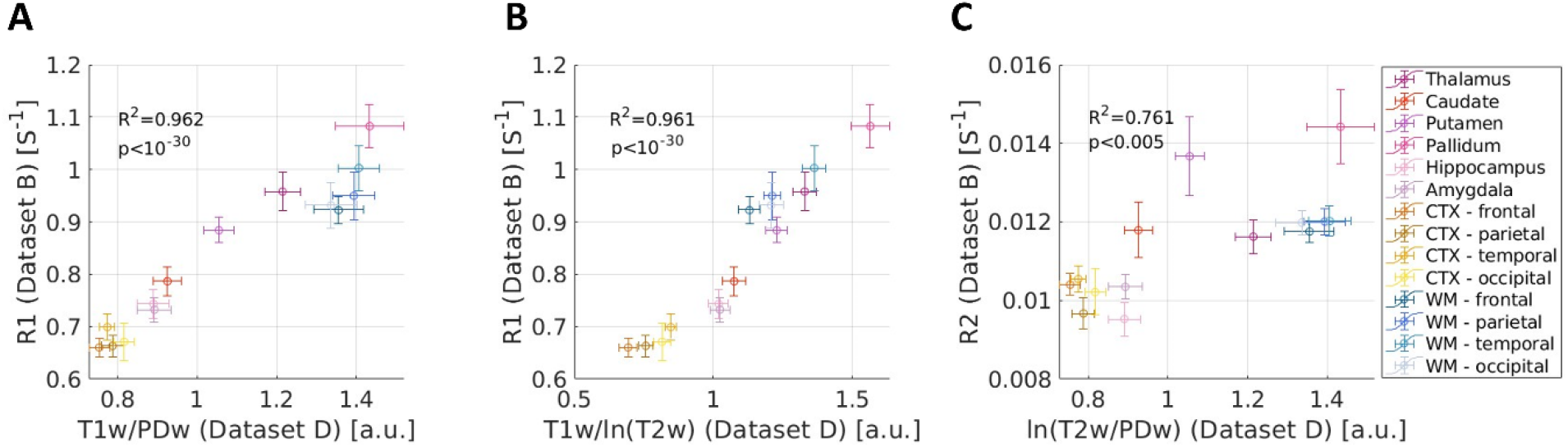
The semi-quantitative quantifiers are robust across datasets. A comparison of quantitative maps from Dataset B (Y-axis, N=22) and semi-quantitative contrasts from Dataset D (X-axis, N=46), across 14 ROIs. **(A)** R1 and T1w/PDw; **(B)** R1 and T1w/ln(T2w); and **(C)** R2 and ln(T2w/PDw). R^2^ values represent Pearson’s correlation coefficient.

Figures

## Discussion

qMRI offers a reliable biophysical interpretation of microstructural changes in the human brain^39^. Unfortunately, in clinical settings the use of qMRI is still rare, which limits quantitative comparisons and the interpretation of such neuroimaging datasets. In this work we test three quantifiers, which are calculated from weighted images commonly used in clinical settings. We show that specific quantifiers resemble qMRI metrics and enhance the applicability of clinical datasets for quantitative interpretation.

The foundational theory of MRI relaxation (which inspired this research) is well established^8,40,41^. Given the known signal equations for single-exponent R1 and R2, our efforts centered on uncovering the latent potential of weighted images to estimate the relaxation rates. We propose a novel way to utilize existing weighted data for quantitative approximations. First, we showed that the observed R1 can be approximated using the T1w/PDw quantifier. We show this using synthetic data, *in vitro* phantom data, and *in vivo* human brain data.

In radiological practice, T1w images are used to visualize the R1 contrast within the subject’s individual brain. A major benefit of T1w/PDw over T1w is that it normalizes the image values across subjects, which allows it to better represent the R1 differences *between* subjects. Hence, this normalization likely removes biases that are common to both images^1,11^.

Another benefit of T1w/PDw is that it allows for the approximation of a “pure” T1-weighted image, reducing PD influences. This rationale also was proposed in Noth et al.^42^, where an MPRAGE image was acquired along with a PDw image. In contrast to that earlier work, the focus of the current work is to combine T1w and PDw images that already exist.

We acknowledge that the R1 signal also depends on the repetition time (TR) and the flip angle (FA), as shown in Eq. 1. However, since most datasets use a specific TR and FA, we simulated the data using unique values that are common in the field. Differences in scanning parameters between and within datasets are likely to shift the observed values. Such contributions also may account for the fact that T1w/PDw is not a perfect linear function of R1, but rather is approximately linear in the range of relevant values across the gray- and white-matter regions^1^.

Furthermore, the equation on which we base this analysis (Eq. 1) is a model from the SPGR equations. This equation is by definition different from the MPRAGE equation^42^, but since both are used with scanning parameters to derive the T1 weighted image, and since both have an inherent PD contribution, the same logic applied for the SPGR T1w image can be generalized also to the MPRAGE T1w datasets (Fig. 3).

Next, we showed that R2 was best approximated by ln(T2w/PDw). Replicating previous study^30^, we found this result across all datasets (synthetic, phantoms, and human datasets). We test whether using the *ln* operation (according to the R2 single-exponent signal equation) with the minimal two echo time images (i.e., T2w, PDw) will allow for a more accurate approximation of R2. Interestingly, this approximation seems to be insensitive to the sequences that were used to produce the PDw and T2w images (Supplementary Fig S1).

In current practice, T1w/T2w has become a standard in large dataset repositories (HCP (https://www.humanconnectome.org/), ABCD). In these studies, it has been proposed as a marker for myelin^11,13,15–17,43^. The interpretation of the T1w/T2w quantifier is still debated^10^; however, since it has become a standard quantifier, both T1w and T2w images are commonly found. Here we test for the best way to use T1w and T2w images to approximate qMRI relaxation. We observed that when T1w/T2w is modified to be T1w/ln(T2w), it can better approximate R1 (Fig 5C). Similarly, ln(T1w/T2w) was a better estimate of R2 than T1w/T2w itself (Fig 5E). These nuanced adjustments hold significant potential for researchers using T1w/T2w, particularly in datasets (e.g., HCP, ABCD) when these are the only available weighted images (and there is no PDw image).

A potential limitation of dividing T1w by ln(T2w) is the fact that it doesn’t cancel out the shared bias between the original images. However, this limitation could potentially be addressed using a postprocessing image bias correction^44^.

Not all clinical datasets have T2w images, but many do contain diffusion MRI sequences (e.g., PNC^45^, ISBI 2017 (https://openneuro.org/datasets/ds002738), Multimodal DTI of the Human Brain (https://openneuro.org/datasets/ds003027/versions/1.2.0), UK-bioBank). We showed that the approximation of R1 also can be done using the T2w image that was acquired as part of the diffusion MRI sequence. Thus, our method can expand the semi-quantitative approximation to additional datasets.

Finally, since we aimed to utilize available clinical datasets for quantitative MRI analysis, we tested the quantifiers on a clinical dataset from the PPMI^38^. We find the semi-quantitative contrasts in the PPMI data correlated with the quantitative maps of Dataset B, as measured across multiple brain ROIs, suggesting our approach to be robust across datasets.

Some of the limitations that we faced stemmed from our decision to work with pre-existing datasets. One limitation of the comparison between quantitative and semi-quantitative measurements in Datasets B is the fact that T2w and PDw are used both to estimate the R2 map and to calculate ln(T2w/PDw). This creates dependencies in the data, which means that correlating between these images and R2 has an inherent bias. Another limitation is that the T1w, T2w, and PDw images were not scanned at the same resolution, requiring down-sampling of T1w and R1. These two limitations were addressed when we acquired Dataset C for replication. However, Dataset C has a small sample size (N=3), which limits the statistical inference.

Additional potential limitations are related to the transmit field (B1+). Specifically, when we divided T1w by PDw or T2w to produce T1w/PDw and ln(T2w/PDw), the transmit field cannot be accounted for, and may not be removed in the division process. However, this bias can be addressed using a non-specific bias correction, e.g., the ANTs biascorrection^34,35^ . Aiming to minimize such biases, in Supplementary Fig. S1,2 we show such example.

In this paper, we show ln(T2w/PDw) can approximate R2. This log linear method had been previously suggested as a quick way to measure R2, and was shown to be highly affected by iron concentrations^30^ and other multi-exponential R2 influences^31^. Naturally, in this paper we compare only semi-quantitative values to single-exponent R1 and R2; however, multi-exponential relaxation processes may contribute to the signal as well^33^. As we acknowledge that comprehensive quantitative mapping can be possible only using multi-exponential mapping, here we highlight the semi-quantitative usage of the existing weighted images, in a simple and fast analysis to approximate the single exponent observed R1 and R2. This method can allow researchers to explore the possibilities available for brain quantifications within existing datasets, where the scanning times and sequences are fixed.

## Conclusions

Weighted images are commonly used in clinical settings and found in many open-source datasets (such as PPMI, ADNI, UK-biobank and ABCD) that rarely contain quantitative MRI mapping. To bridge this gap, we tested three quantifiers, based on simple combination of weighted images. We found that T1w/PDw yields a satisfactory approximation of R1, and rectifies image biases. Similarly, ln(T2w/PDw) is an accurate approximation of R2. We also refined the well-accepted T1w/T2w approach, offering better quantitative approximations (ln(T1w/T2w), T1w/ln(T2w)), and extended the analysis to databases with diffusion MRI data. When applied to a clinical dataset, we showed substantive correspondence between semi-quantitative contrasts and quantitative maps. Our work, therefore, presents a pathway for harnessing widely available weighted images to offer approximate quantitative insights, paving the way for enhanced neuroimaging analyses in both research and clinical settings.

## Methods

### Theory

In this section, we follow the MRI signal equations to identify quantifiers that can be approximated from weighted images.

The typical T1w signal *S* can be generated using a spoiled gradient echo (SPGR) sequence^46^:

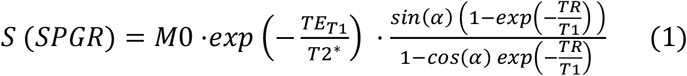

where α is the flip angle, M0 is the magnetization at equilibrium, TR is the repetition time, and *TE*_*T*1_ is the echo time for the T1 signal.

For short echoes, we can assume 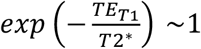.

The typical T2w and PDw signals can be generated using a spin echo (SE) sequence^1^:

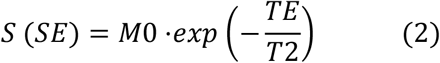

By varying the echo time (TE), one can generate the PDw signal (short TE) and the T2w signal (long TE).

In both Eq. 1. and Eq. 2, the PDw contrast dominates the M0 parameter (*M*0 ≈ *PD* . *G*, where G is a scale factor that characterizes the receive-coil inhomogeneity gain^11^). When we divide Eq. 1 by the PDw contributions, we get a pure T1w signal. Hence, we predict that 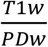 will provide a better approximation of the R1 contrast:

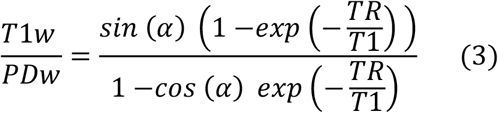

Note that the signal equations for PDw and T2w differ only by the TE. Since PDw entails a short TE, 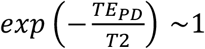, so T1w/PDw will not contain PD influences. On the other hand, since T2w has a long TE, 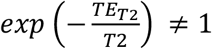, and so the contrast T1w/T2w will be a mix of both T1 and T2 processes.

We note that T1w also can be generated using other sequences (for example, MPRAGE^47^). However, since T1w images typically contain a T1-weighting term that is multiplied by a PD-weighting term, the same logic can be applied.

A similar heuristic can be applied for Eq. 2 (keeping in mind that both PDw and T2w derive from the same sequence):

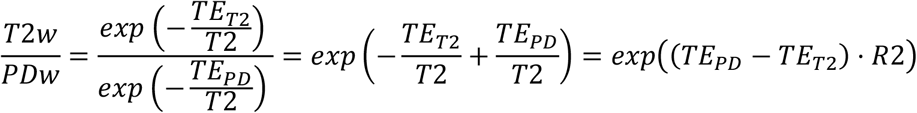

We denote 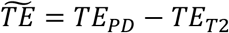. We apply the natural log (ln) on 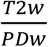 to obtain a linear function of R2 (also known as log-linear method^30^):

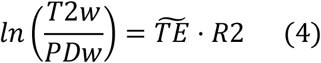

So, to achieve a direct correlation with R2 we divide In 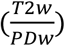 by 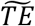.

Thus, by these mathematical operations, we can test two quantifiers: T1w/PDw, which approximates R1 (Eq. 3); and ln(T2w/PDw), which approximates R2 (Eq. 4).

## Datasets

In order to test the semi quantitative quantifiers, we examined four datasets. Three of them (A, B, and D) were pre-existing, and dataset C was collected as a validation. Datasets A, B, and C were collected at the same scanner at the Hebrew University by our group, while dataset D was collected on multiple scanners elsewhere.

All data preprocessing and analysis were preformed using MATLAB^48^.

### Dataset A - Phantom data

The lipid dataset was designed and collected in a previous work by our group^49^. In short, four types of lipid mixtures were produced (phosphatidylcholine – PC; phosphatidylcholine and cholesterol – PC:chol; phosphatidylserine – ps; and sphingomyelin – spg), at different concentrations. All samples were scanned using a 3T Siemens MAGNETOM Skyra scanner equipped with a 32-channel head receive-only coil. Data were collected at the Edmond and Lily Safra Center’s Neuroimaging Unit at the Hebrew University of Jerusalem, Israel.

### Data acquisition

For R1 and PD mapping, 3D images were acquired using a fast low angle shot (FLASH) sequence with four flip angles (α = 4°, 8°, 16°, 30°), TE = 3.91 ms, TR = 18 ms, field of view (FoV) of 205 mm^2^, and voxel size 1.1 mm × 1.1 mm × 0.9 mm. For bias estimation, single-slice images were acquired using a spin echo inversion recovery (SEIR) sequence with an adiabatic inversion pulse and inversion times of TI = 2000, 1200, 800, 400, 50 ms, TR = 2540 ms, TE = 73 ms, FoV of 222 mm^2^, and voxel size of 1.2 mm × 1.2 mm × 2.0 mm.

For R2 mapping, images were acquired with multi-spin-echo sequences with 15 equally spaced echo times between TE = 10.5 and TE=157.5 ms, TR = 4940 ms, FoV of 207 mm^2^, and voxel size of 2 mm isotropic.

The T1w values used SPGR scans with α=30°. The shortest and longest TEs for the multi-spin-echo scans (TE= 10.5 and 157.5 ms) were used for the PDw and T2w values, respectively.

### Estimation of qMRI parameters and analysis

The R1 map was estimated using the variable flip angle approach. The R1 and PD maps, as well as the B1+ excitation bias, were computed using mrQ software^50,51^. The PD values for all lipids were normalized to the value of the water in the cuvette. The mrQ software was modified to suit the phantom system: since the agar-Gd filling the box around the samples is homogeneous, it therefore can be assumed to have a constant R1 value. The R2 map was estimated from a multi-spin-echo sequence using the echo-modulation curve (EMC) algorithm^52^. We calculated the T1w/T2w and T1w/PDw values using voxel-to-voxel division of the contrasts’ values. For ln(T2w/PDw), we divided the contrasts’ values, and then derived the natural logarithm, and multiplied the result by 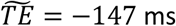. All phantom data underwent bias correction using a 3^rd^-degree polynomial over the agar mask, as was done in a previous study^53^.

### Simulated data

To examine our approach from a theoretical perspective, we produced simulated data. Simulated weighted data was produced for each cuvette by plugging the quantitative values (i.e., R1 and R2 simulated values in the range of real R1 and R2 values) into the relevant signal equations. For the synthetic T1w images, we use Eq. 1 and assume 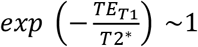, α = 30°, and *TR* = 18 *ms*. For synthetic T2w images, we use Eq. 2 with *TE*_*T*2_ = 157.5 *ms*. We assume that the simulated PDw contrast can be best represented by Eq. 2 given a short TE (*TE*_*PD*_ = 10.5 *ms*), so that 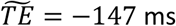.

### Dataset B - HUJI dataset 1

This human dataset was collected by Filo et al. ^53^, using the same scanner as in Dataset A.

#### Participants

From Filo et al.’s original dataset of 41 subjects^9^, we selected 22 individuals who had MPRAGE T1w scans, since these scans are considered to be the gold standard for T1w scans in clinical settings^54^. The original study focused on aging, and thus our data contained two age groups: younger adults (n=13, age 26.7±2.4 years, 7 females) and older adults (n=9, age 67.1±5.5 years, 2 females). Of the 22 subjects, 20 also had B0 diffusion images. All study procedures were approved by the Helsinki Ethics Committee from Hadassah Hospital, Jerusalem, Israel. Written informed consent was obtained from all participants before the procedure.

#### Data acquisition

For quantitative R1 and PD mapping, 3D SPGR echo images were acquired with different flip angles (α = 4°, 10°, 20°, and 30°). Each image included five equally spaced echoes (TE = 3.34– 14.02 ms) and the TR was 19 ms (except for six younger subjects, for whom the scan included only one TE = 3.34 ms). The scan resolution was 1 mm isotropic. For calibration, an additional spin-echo inversion recovery (SEIR) scan was acquired with an echo-planar imaging (EPI) read-out (SEIR-epi). This scan was done with a slab-inversion pulse and spatial-spectral fat suppression. For SEIR-epi, the TE/TR were 49/2920 ms. The TI were 200, 400, 1200, and 2400 ms. We used 2-mm in-plane resolution with a slice thickness of 3 mm. The SEIR-epi read-out was performed using 2× acceleration. For quantitative R2 mapping, images were acquired with a multi spin-echo sequence with 15 equally spaced spin echoes between 10.5 and 157.5 ms. The TR was 4.94 s. The scan resolution was 1.2 mm isotropic. For the diffusion B0 images, whole-brain DTI measurements were performed using a diffusion-weighted spin-echo EPI sequence with a resolution of 1.5 mm isotropic. Diffusion weighting gradients were applied at 64 directions, and the strength of the diffusion weighting was set to b = 2000 s/mm^2^ (TE/TR = 95.80/6000 ms, G = 45mT/m, δ = 32.25 ms, Δ = 52.02 ms). The data include eight non-diffusion-weighted images (b = 0). For T1w values, we used MPRAGE scans with a resolution of 1 mm isotopic. The shortest and longest TEs for the multi-spin-echo scans (TE=10.5 and 157.5 ms) were used for the PDw and T2w values, respectively.

#### Estimation of qMRI parameters and analysis

The PD and R1 estimations were computed using the mrQ^50,51^ and Vista Lab software packages. The R2 map was estimated from a multi-spin-echo sequence using the EMC algorithm^52^. Diffusion analysis was done using the FDT toolbox in FSL^36,55^. We used rigid-body alignment to register the diffusion MRI (dMRI) data to the imaging space of R1 (Flirt, FSL ^36,55^). All quantifiers (T1w/PDw, T1w/T2w, and ln(T2w/PDw)) were computed by voxel-to-voxel division. We calculated the ln(T2w/PDw) quantifier using 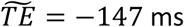 (as shown in Eq. 4).

#### Brain registration and segmentation

We registered the dMRI, qMRI and weighted images to the lower-resolution R2 space (2 mm^3^ isotropic). We used the SynthStrip toolbox^56^ to create a brain mask. We then applied rigid-body registration using SPM 12 (https://www.fil.ion.ucl.ac.uk/spm/software/spm12/), with the lower-resolution R1 map as a reference.

Brain segmentation was done using FreeSurfer^57^, with the T1w image as a reference. All steps were inspected by eye to ensure image quality.

ANTs bias correction^34,35^ was applied to the T1w, T2w, and PDw images independently, and then all quantifiers were computed voxel-to-voxel as described previously.

Since the ANTs bias correction algorithm tends to shift the overall values of the images, for each corrected image we calculated the ratio between the pre- and post-correction image medians, and multiplied by this ratio to shift the values back to their original range.

### Dataset C – HUJI dataset 2 (validation dataset)

In this dataset, we acquired scans for weighted images and scans for quantitative mappings independently, using a separate sequence for each map or weighted image. We collected the data using the same scanner used for datasets A and B, to serve as a validation dataset designed specifically for this study (unlike datasets A, B, and D, which were pre-existing).

#### Participants

Three individuals (age 28.0±1.5 years, one female) were scanned. All study procedures were approved by the Helsinki Ethics Committee from Hadassah Hospital, Jerusalem, Israel. A written informed consent was obtained from all participants before the procedure.

#### Data acquisition

For quantitative R1 and PD mapping, 3D SPGR echo images were acquired with different flip angles (α = 4°, 10°, 20°, and 30°). The TE/TR were 8.68/19 ms. The scan resolution was 2 mm isotropic. For calibration, we acquired an additional SEIR-epi scan. This scan was done with a slab-inversion pulse and spatial-spectral fat suppression. The TE/TR were 49/2920 ms, with an adiabatic inversion pulse and inversion times of TI = 2400, 1200, 400, and 200. The scan resolution was 2 mm isotropic.

For quantitative R2 mapping, images were acquired with a multi spin-echo sequence with 10 equally spaced spin echoes between 12 and 120 ms. The TR was 4210 ms. The scan resolution was 2 mm isotropic.

For the T1w image, we scanned an MPRAGE sequence with a resolution of 2 mm isotropic. For the T2w images, we used a SIEMENS T2w structural scan with TE/TR of 482/3200 ms and a resolution of 2 mm isotropic. For the PDw images, we scanned a SIEMENS T2 scan with a single echo time, with TE/TR of 12/4210 ms and a resolution of 2 mm isotropic.

#### Estimation of qMRI parameters

The PD and R1 estimations were computed using the mrQ^50,51^ and Vista Lab software packages.

The R2 map was estimated from a multi-spin-echo sequence using the EMC algorithm^52^.

The semi-quantitative quantifiers (T1w/PDw, T1w/T2w, and ln(T2w/PDw)) were computed by voxel-to-voxel division. We calculated the ln(T2w/PDw) using 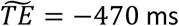 (as in Eq. 4).

#### Brain registration and segmentation

Image registration, segmentation, and correction was done as described in Dataset B.

### Dataset D - PPMI

#### Participants and data

For our fourth dataset, we used data from an open-source dataset of the Parkinson’s Progression Marker Initiative (PPMI). We used a subset of the dataset, which was selected in a previous published work by our group^23^. Our dataset consists of 46 healthy control subjects (age 65±6 years, 17 females).

#### Data acquisition

MRI data were collected using 3T SIEMENS Trio scanners, in 8 different sites. T1w images were acquired using an MPRAGE generalized autocalibrating partially parallel acquisitions (MPRAGE-GRAPPA) sequence, with a sagittal slice thickness of 1 mm and an in-plane resolution of 1 mm^2^, and the flip angle was α = 9°. The T2w and PDw images were acquired using a turbo spin echo (TSE) sequence with an axial slice thickness of 3 mm and an in-plane resolution of 0.94-mm^2^, with TR=3000 ms and TE=11 ms (for T2w) or 101 ms (for PDw).

#### Brain registration and segmentation

We registered, aligned, and resampled the T2w and PDw images to the T1w image space, using software packages from SPM and MRtrix3 (www.mrtrix.org/). All steps were inspected by eye to ensure image quality. Segmentation was done in the same procedure as in Datasets B and C^57^.

### Statistical analysis

All statistical analyses were performed using MATLAB^48^.

Correlations between quantifiers were calculated using Pearson’s correlation.

To quantify the differences between the correlations, we performed cross-validation for the following pairs of variables: R1 and T1w; R1 and T1w/T2w; R1 and T1w/PDw; R2 and R2w; R2 and T1w/T2w; and R2 and ln(T2w/PDw), for Dataset B. For each of the pairs we randomly selected 90% of the voxels and used them to produce a linear fit, which we tested on the remaining 10%. We then calculated the root mean square error (RMSE):

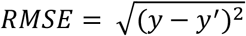

where *y* is the real quantifier value, and *y’* is the predicted quantifier value. We repeated this process 1,000 times for each pair, and then averaged the RMSE over these 1,000 repetitions.

## Data Availability

All data produced in the present study are available upon reasonable request to the authors

https://www.ppmi-info.org/access-data-specimens/download-data

