## Supplemental Material for "Approximating R1 and R2: a quantitative approach to clinical weighted MRI"

### Supplementary material

#### Figure S1

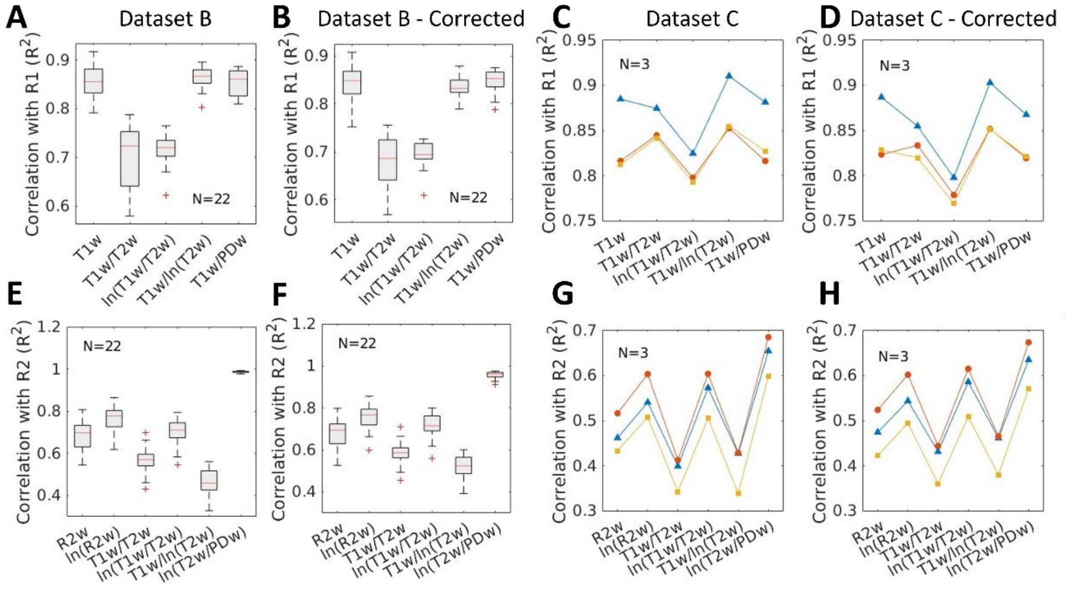


**Fig. S1 – R1 and R2 correlations to weighted quantifiers across subjects**

A comparison of R1 and R2 to different quantifiers derived from weighted images, across subjects. **(A)** Group correlations between R1 and five quantifiers. Each box represents the single-subject correlation coefficients across 22 subjects (Dataset B). Significant differences are found with T1w, T1w/T2w, and ln(T1w/T2w), but not with T1w/ln(T2w) or T1w/PDw. **(B)** Same as **(A)**, but for quantifiers calculated from bias-corrected images. **(C)** Individual correlations between R1 and five quantifiers for three subjects in Dataset C. Each color represents a single-subject correlation coefficient, connecting lines are for visual assistance and do not contain additional information. T1w/ln(T2w) yields the highest significant correlation across subjects. **(D)** Same as **(C)**, but for quantifiers calculated from bias-corrected images. **(E-H)** same as **(A-D)**, respectively, but for correlations of R2 with six quantifiers.

### Figure S2

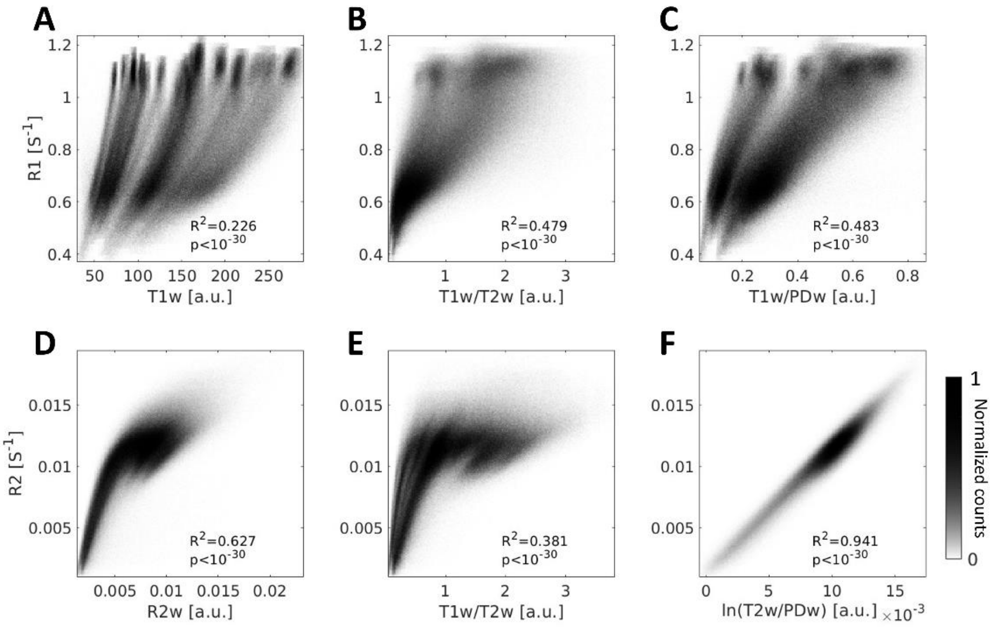


**Fig. S2 - R1 and R2 can be approximated using the T1w/PDw and ln(T2w/PDw) quantifiers in human data *in vivo* in bias-corrected images**

A comparison of R1 and R2 to different quantifiers derived from bias-corrected weighted images. **(A-C)** Voxel-wise 2D-histograms of R1 values with either **(A)** T1w; **(B)** T1w/T2w; or **(C)** T1w/PDw. **(D-F)** Voxel-wise 2D-histograms of R2 voxels with either **(D)** R2 **(C)** T1w/T2w; or **(F)** ln(T2w/PDw). The ln(T2w/PDw) quantifier shows the highest correlation with R2. $R^2$ represents Pearson's correlation coefficients. Data pooled from Dataset B (N=22). Colorbar represents normalized voxel counts.

### Figure S3

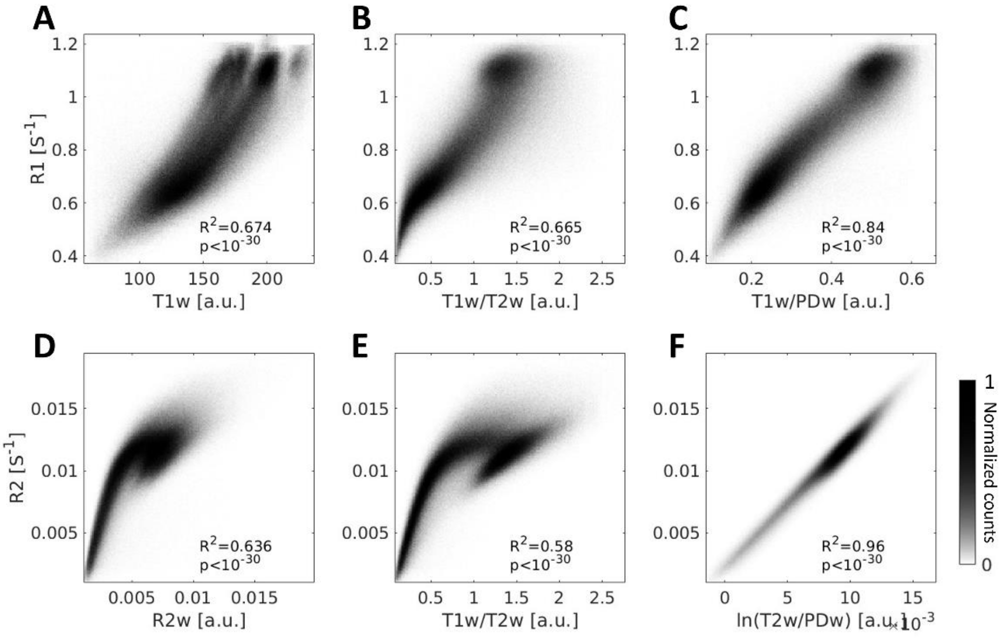


**Fig. S3 - R1 and R2 can be approximated using the T1w/PDw and ln(T2w/PDw) quantifiers in human data *in vivo* in bias-corrected and normalized images**

Same as Fig. S2, except with the addition of image normalization to the process. As in Fig. S2, the highest correlations were found for T1w/PDw (with R1) and ln(T2w/PDw) (for R2).

### Figure S4

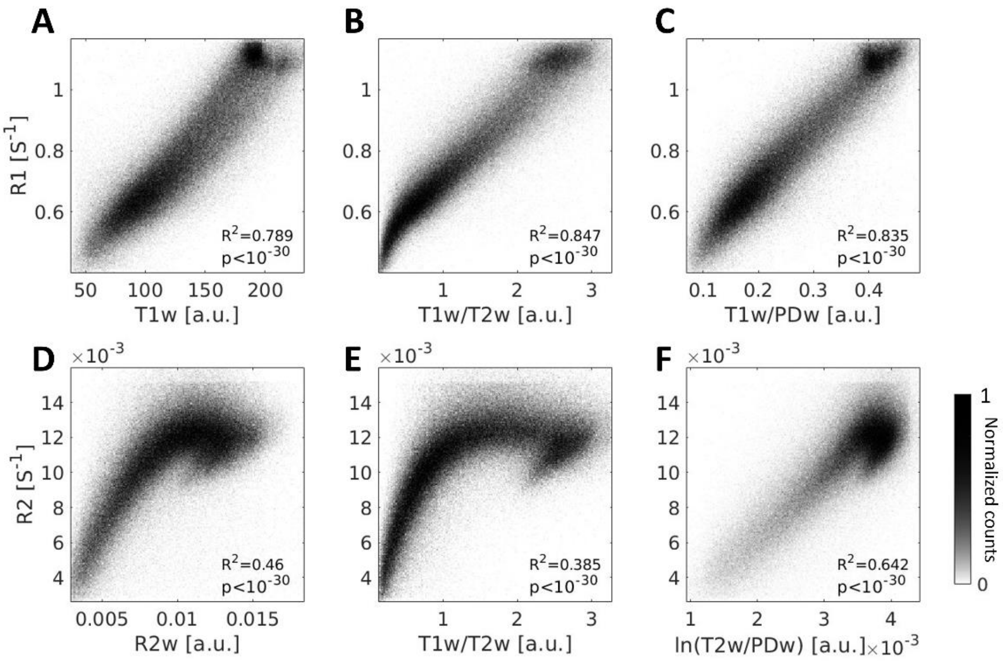


**Fig. S4 – R1 and R2 can be approximated using the T1w/PDw and ln(T2w/PDw) quantifiers in human data *in vivo***

A comparison of R1 and R2 to different quantifiers derived from weighted images. This figure is similar to Figure 3 in the main text, but for a different dataset (Dataset C; N=3). **(A-C)** Voxel-wise 2D-histograms of R1 voxels pooled from Dataset C (N=3) with either **(A)** T1w; **(B)** T1w/T2w; or **(C)** T1w/PDw. The T1w/T2w and T1w/PDw and quantifiers shows the highest correlation with R1, and are not significantly different from each other. **(D-F)** Voxel-wise 2D-histograms of R2 voxels pooled from Dataset C (N=3) with either **(D)** R2w; **(E)** T1w/T2w; or **(F)** ln(T2w/PDw). The ln(T2w/PDw) quantifier shows the highest correlation with R2. $R^2$ represents Pearsons' correlation coefficients. Colorbar represents normalized voxel counts.

### Figure S5

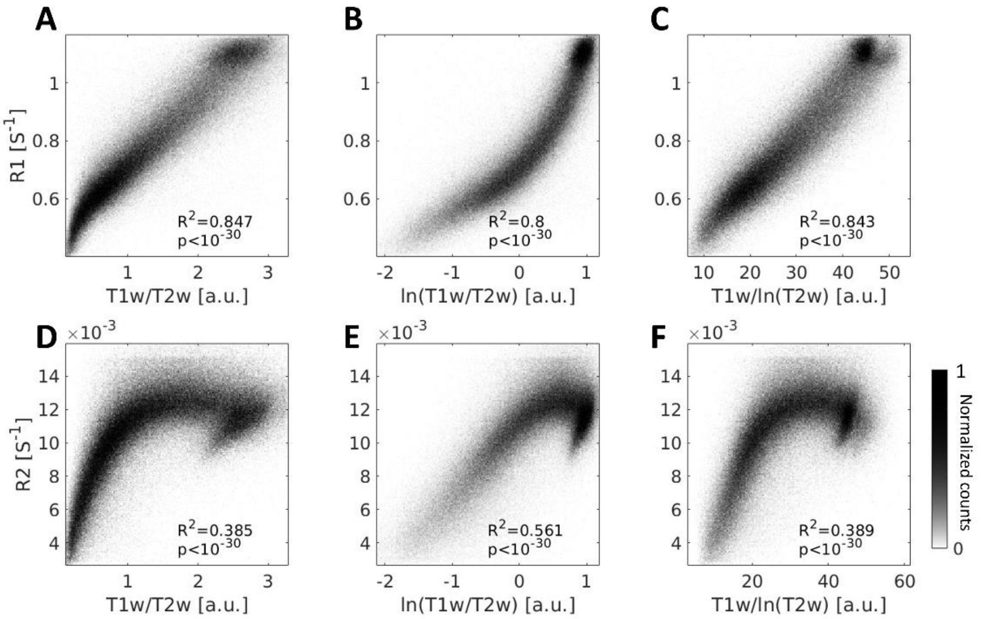


**Fig. S5 – Adjusting the T1w/T2w quantifier improves the approximation of R1 and R2**
A comparison of R1 and R2 to different quantifiers derived from weighted images, across subjects. This figure is similar to Figure 4 in the main text, but for a different dataset (Dataset C; N=3).
**(A-C)** Voxel-wise 2D-histograms of R1 voxels pooled from all subjects (y-axis), against either (**A**) T1w/T2w; (**B**) ln(T1w/T2w); or (**C**) T1w/ln(T2w). The results differ from those of Dataset B in that T1w/ln(T2w) is as good as the T1w/T2w approximation of R1. **(D-F)** The same as **(A-C)** but for R2. Similar to Figure 5, ln(T1w/T2w) improves the T1w/T2w approximation of R1. $R^2$ represents Pearson's correlation coefficients. Colorbar represents normalized voxel counts.
